# Effect of Beta Blockers on Exercise Capacity, Diastolic Function, and Quality of Life in Patients with Heart Failure with Preserved Ejection Fraction: A Secondary Analysis of INDIE-HFpEF and RELAX

**DOI:** 10.1101/2024.09.20.24314104

**Authors:** Frank R. Weigel, Alexandria Miller, Vaiibhav Patel, Garrie Haas, Sakima Smith

## Abstract

**Background:** The use of beta blockers in patients with heart failure with preserved ejection fraction (HFpEF) is common, with about 75% of patients in recent landmark clinical trials on beta blockers. Though the implementation of this medication class is routine, there is sparse data to support their use. Furthermore, beta blocker effects on exercise capacity, diastolic function, and quality of life in HFpEF patients is unclear.

**Methods:** A retrospective cohort study was completed using patient-level data from two prior randomized trials. Cohorts were generated based on beta blocker use at the time of trial enrollment, demographic information was compared. Primary outcomes assessed were exercise capacity, diastolic function, and quality of life metrics. The results of baseline testing were utilized to avoid potential bias from each trial’s intervention.

**Results:** After multivariate linear regression, HFpEF patients on beta blockers had no difference in exercise capacity (peak VO2 (mL/kg/min): 12.5 vs 13.5, P=0.933), diastolic function (average E/e’: 16 vs 14, P=0.125; left atrial volume index (mL/m2): 47 vs 42, P=0.665; peak tricuspid regurgitation velocity (m/s): 2.85 vs 2.70, P=0.165), or quality of life survey scores (KCCQ: 54 vs 60, P=0.206; MLHFQ 44 vs 48, P=0.762) compared to those not taking beta blockers.

**Conclusions:** In this secondary analysis of patient-level data, there was no association with beta blocker use and worsened exercise capacity, diastolic function, or quality of life in HFpEF patients. Randomized controlled trials are necessary to definitively determine the clinical and functional impact of beta blockers in HFpEF.

**RESEARCH PERSPECTIVE:**

- In this secondary analysis or INDIE-HFpEF and RELAX, there was no significant association with beta blocker use and worsened exercise capacity, diastolic dysfunction, or quality of life in HFpEF patients.
- With little available evidence to suggest clinical or functional benefits from implementation of beta blockers in HFpEF, randomized controlled trials are warranted to more definitively evaluate the potential effects of this medication class.
- Evaluating unique HFpEF phenotypes with different comorbidity profiles would provide specific insights that could be translated to the clinical management of HFpEF patients.

## INTRODUCTION

Over 6.5 million Americans over the age of 20 years have heart failure (HF), with a trend toward increasing prevalence of heart failure with preserved ejection fraction (HFpEF).^1^ In the population of patients with HFpEF, the use of beta blockers is common despite little evidence to support improved clinical outcomes with this medication class. In recent landmark clinical trials, a majority of HFpEF patients were using beta blockers at trial enrollment.^2–5^ Even with the widespread use of beta blockers in HFpEF patients, recent HF guidelines do not offer insight into the best practices or clinical considerations for managing these medications in this population.^6^ With multiple observational and secondary analyses reporting conflicting findings, the effects that beta blockers have on morbidity and mortality in patients with HFpEF remain unclear.

Proposed benefits from beta blocker use in HFpEF are by reducing the degree of sympathetic hyperactivity that is known to occur in patients with HF, regardless of left ventricular ejection fraction (LVEF), as well as blunting the rate of development of diastolic dysfunction by preventing myocardial fibrosis.^7^ However, the negative chronotropic effect of beta blockers is thought to limit HFpEF patients from significantly augmenting cardiac output in times of stress or exertion, as these patients commonly are unable to increase stroke volume due to non-compliant left ventricles.^8^ This effect of beta blockers is likely to worsen exertional symptoms in patients with HFpEF, as their underlying hemodynamic profiles limit the ability to compensate for stressed blood volumes leading to pulmonary vasculature congestion.^9^

The only randomized clinical trial (n=245) to directly assess the impact of beta blocker use in HFpEF patients demonstrated no difference in cardiovascular death or HF hospitalization compared to placebo.^10^ In a network meta-analysis examining the therapeutic effects of different classes of medications in patients with HF and LVEF above 40%, beta blockers did not have a significant association with rates of HF hospitalizations, cardiovascular death, or all-cause death.^11^ One recent observational study of registry data found an association with beta blocker use and higher rates of heart failure hospitalizations in patients with HFpEF, with higher risk as LVEF increased.^12^ Similar findings were reported from a secondary analysis of TOPCAT, where beta blocker use was associated with increased risk of HF hospitalization but not cardiovascular mortality.^13^

Beyond adverse clinical outcomes, exercise intolerance and dyspnea are common symptoms that lead to poor quality of life and functional status in patients with HFpEF. One crossover clinical trial, PRESERVE-HR, examined the effects of beta blocker discontinuation in HFpEF patients with chronotropic incompetence. It was shown that beta blocker withdrawal improved maximal functional capacity and quality of life in this patient population.^14^ The potential effects of beta blockers on exercise capacity and diastolic function in HFpEF patients without chronotropic incompetence is still relatively unknown. Hence, the goal of this study was to examine the effects of beta blocker use on exercise capacity, echocardiographic parameters of diastolic function, and quality of life in HFpEF patients.

## METHODS

A retrospective observational cohort study was conducted. This study was considered exempt by the institutional review board at The Ohio State University Wexner Medical Center in Columbus, OH, USA. Analyses were performed on deidentified patient data from the INDIE-HFpEF [Effect of Inorganic Nitrite vs Placebo on Exercise Capacity Among Patients With Heart Failure With Preserved Ejection Fraction: The INDIE-HFpEF Randomized Clinical Trial] and RELAX [Effect of Phosphodiesterase-5 Inhibition on Exercise Capacity and Clinical Status in Heart Failure with Preserved Ejection Fraction: a Randomized Clinical Trial] trials as obtained through the Biologic Specimen and Data Repository Information Coordinating Center of the National Institutes of Health.^15^ Two populations of patients were created based on beta blocker use at time of trial enrollment, and baseline demographics were compared between populations. Outcomes were assessed based upon baseline exercise, echocardiographic, and survey data to avoid potential confounding from the effects of each study’s intervention.

INDIE-HFpEF, published in 2018, was a double-blind, placebo-controlled, crossover trial of 105 patients with HFpEF across 17 trial sites in the United States. The objective of the study was to determine the effect of inhaled, nebulized inorganic nitrite on exercise capacity. Key inclusion criteria were LVEF ≥ 50% and objective evidence of HF, as demonstrated by previous HF hospitalization with imaging evidence of pulmonary congestion, elevated left ventricular (LV) end-diastolic pressure and/or pulmonary capillary wedge pressure at rest (≥ 15 mmHg) or with exercise (≥ 25 mmHg), elevated BNP (> 200 pg/mL) and/or NT-proBNP (≥ 400 pg/mL), or echocardiographic evidence of diastolic dysfunction (medial E/e’ ratio ≥ 15 or left atrial enlargement) with chronic loop diuretic treatment. During the nitrite phase, there was no significant difference in mean peak oxygen consumption (peak VO2), daily activity levels, Kansas City Cardiomyopathy Questionnaire (KCCQ) clinical summary score, echocardiographic E/e’ ratio, or N-terminal pro-brain natriuretic peptide (NT-proBNP).^16^

RELAX, published in 2013, was a double-blind, placebo-controlled, randomized clinical trial of 216 patients with HFpEF across 26 trial sites in the United States and Canada. The objective of the study was to determine the effect of phosphodiesterase-5 inhibitors on exercise capacity and clinical status. Key inclusion criteria were LVEF ≥ 50%, New York Heart Association (NYHA) functional class II-IV symptoms on stable medical therapy, and objective evidence of heart failure (prior HF hospitalization, acute HF therapy with intravenous diuretic therapy, chronic loop diuretic therapy for HF with left atrial enlargement, or invasively documented elevation in left ventricular filling pressures). Compared with placebo, there was no significant difference in median change in peak oxygen consumption, change in six-minute walk test (6MWT) distance, or mean clinical status rank score.^17^ In both trials, enrolled patients underwent baseline evaluation that included demographics, medical history, medication review, laboratory studies, cardiopulmonary exercise testing, and echocardiogram.

Three primary outcomes were assessed in this study using multiple parameters. Exercise capacity was assessed by peak VO2, peak respiratory exchange ratio (RER), ventilatory efficiency (VE/VCO2), 6MWT, and peak heart rate (HR). Diastolic function was assessed by 6 echocardiographic parameters: average E/e’, lateral e’ velocity, medial e’ velocity, left atrial volume index (LAVI), peak tricuspid regurgitation (TR) velocity, and pulmonary artery systolic pressure (PASP). Quality of life was assessed by patient responses to validated symptom surveys, KCCQ and Minnesota Living with Heart Failure Questionnaire (MLHFQ). Secondary outcomes assessed were baseline NT-proBNP, as well as HF hospitalizations and death during trial follow-up period.

Categorical variables were summarized as frequencies (n) and percentages (%) and compared using Pearson’s Chi-squared test or Fisher’s exact test, as appropriate. Continuous variables were summarized as median and inter-quartile ranges (IQR) and compared using Wilcoxon rank sum test. Stepwise multivariable linear regression models were used to investigate the association between continuous outcomes and beta blocker use. In the stepwise multivariable linear regression analysis, we considered a set of potential confounders, including demographic information (age, sex, body mass index (BMI)), clinical parameters (atrial fibrillation, myocardial infarction, chronic obstructive pulmonary disease, diabetes mellitus, hypertension, serum creatinine), and NYHA functional class. The statistical analysis was performed using R software (version 4.2.2; R Core Team, R Foundation for Statistical Computing, Vienna, Austria).

## RESULTS

In total, 320 patients were included in the analysis, 216 from RELAX and 104 from INDIE-HFpEF. There were 230 patients using beta blockers at time of trial enrollment (72%). Baseline demographics are shown in Table 1. In general, patients enrolled in these two clinical trials were older (average age 68 years old), white (90%), obese (average BMI 34 kg/m^2^) individuals with an average LVEF of 61% (range 38-77%). Of those studied, 54% of participants were female. In the 12 months prior to trial enrollment 31% of patients had a HF hospitalization, there was no difference in HF hospitalization rates prior to trial enrollment in patients taking beta blockers compared to those not taking beta blockers (34% vs 26%, P = 0.15). At baseline, all patients were either NYHA functional class II (45%) or NYHA functional class III (55%).

**Table 1.** Baseline demographics based upon beta blocker use at trial enrollment.

| Characteristic | No Beta Blocker (90) | Beta Blocker (230) | P Value |
| --- | --- | --- | --- |
| Age (years) | 67 (59, 75) | 69 (63, 77) | 0.067 |
| Sex |  |  | 0.036 |
| Female | 54 (60) | 108 (47) |  |
| Male | 36 (40) | 122 (53) |  |
| Race |  |  | 0.7 |
| White | 82 (91) | 206 (90) |  |
| Black | 7 (8) | 17 (7) |  |
| Other | 1 (1) | 7 (3) |  |
| BMI (kg/m<sup>2</sup>) | 33 (28, 38) | 34 (29, 39) | 0.4 |
| HR (bpm) | 72 (64, 82) | 68 (60, 77) | 0.008 |
| SBP (mmHg) | 124 (115, 136) | 128 (116, 140) | 0.2 |
| DBP (mmHg) | 70 (64, 79) | 70 (63, 80) | > 0.9 |
| LVEF (%) | 60 (58, 66) | 60 (55, 65) | 0.2 |
| NYHA Functional Classification |  |  | 0.5 |
| II | 43 (48) | 101 (44) |  |
| III | 47 (52) | 129 (56) |  |
| Medical History |  |  |  |
| Atrial Fibrillation | 37 (41) | 121 (53) | 0.064 |
| Chronic Obstructive Pulmonary Disease | 16 (18) | 38 (17) | 0.8 |
| Diabetes Mellitus | 28 (31) | 102 (44) | 0.03 |
| Hyperlipidemia | 58 (64) | 178 (77) | 0.018 |
| Hypertension | 62 (69) | 206 (90) | < 0.001 |
| Prior Myocardial Infarction | 5 (6) | 25 (11) | 0.14 |
| HF Hospitalization in Prior 12 Months | 23 (26) | 78 (34) | 0.15 |
| Serum Creatinine | 1.10 (0.83, 1.30) | 1.11 (0.89, 1.43) | 0.13 |
| Medications |  |  |  |
| RAASi | 48 (53) | 160 (70) | 0.006 |
| MRA | 16 (18) | 40 (17) | > 0.9 |
| Loop Diuretic | 61 (68) | 184 (80) | 0.02 |
| Statin | 48 (53) | 155 (67) | 0.019 |
| Anti-Arrhythmic | 10 (11) | 14 (6) | 0.12 |

Median (IQR); n (%)
Abbreviations: BMI, body mass index; kg, kilogram; m, meter; HR, heart rate; bpm, beats per minute; SBP, systolic blood pressure; mmHg, millimeter Mercury; DBP, diastolic blood pressure; LVEF, left ventricular ejection fraction; NYHA, New York Heart Association; HF, heart failure; RAASi, renin angiotensin aldosterone system inhibitor; MRA, mineralocorticoid receptor antagonist

### Baseline Demographics

Baseline demographics, according to beta blocker use, for each individual clinical trial are shown in Supplementary Table 1. Overall, there were notable differences between cohorts. HFpEF patients taking beta blockers were more likely to be male (53% vs 40%, P = 0.036), and patients on beta blockers had lower resting HR (68 bpm vs 72 bpm, P = 0.008). Regarding medical comorbidities, HFpEF patients on beta blockers were more likely to have diabetes mellitus (44% vs 31%, P = 0.03), hyperlipidemia (77% vs 64%, P = 0.018), and hypertension (90% vs 69%, P < 0.001). There were no differences in the rates of atrial fibrillation (53% vs 41%, P = 0.064) or prior myocardial infarction (11% vs 6%, P = 0.14). HFpEF patients on beta blockers were more likely to be concomitantly using renin angiotensin aldosterone system inhibitors (RAASi) (70% vs 53%, P = 0.006), loop diuretics (80% vs 68%, P = 0.02), and statins (67% vs 53%, P = 0.019).

### Exercise Parameters

Outcomes for exercise parameters, echocardiographic parameters, symptom assessments, and select clinical outcomes are shown in Table 2. On initial testing, HFpEF patients on beta blockers had a mean peak VO2 of 12.5 mL/kg/min (SD 3.3) with a mean peak RER of 1.10 (SD 0.1). Patients on beta blockers also had a mean VE/VCO2 of 33 (SD 8), mean 6MWT distance of 303 meters (SD 111), and a mean peak HR of 108 bpm (SD 24). HFpEF patients not on beta blockers had a mean peak VO2 of 13.5 mL/kg/min (SD 3.3) with a mean peak RER of 1.12 (SD 0.1). Patients not on beta blockers also had a mean VE/VCO2 of 33 (SD 6), mean 6MWT distance of 294 meters (SD 110), and a mean peak HR of 122 bpm (SD 25). In both groups, baseline peak oxygen consumption and 6MWT distance were reduced compared to normal standards.

**Table 2.** Outcomes for exercise parameters, echocardiographic parameters, symptom assessments, and select clinical outcomes.

| **Outcome** | **No Beta Blocker (90)** | **Beta Blocker (230)** |
| --- | --- | --- |
| *Exercise Parameters* |  |  |
| Peak VO<sub>2</sub> (mL/kg/min) | 13.5 (3.3) | 12.5 (3.3) |
| Peak RER | 1.12 (0.10) | 1.10 (0.10) |
| VE/VCO<sub>2</sub>* | 33 (6) | 33 (8) |
| 6MWT (m)<sup>†</sup> | 294 (110) | 303 (111) |
| Peak HR | 122 (25) | 108 (24) |
| *Echocardiographic Parameters* |  |  |
| Average E/e' | 14 (8) | 16 (9) |
| Lateral e' (m/s) | 0.09 (0.03) | 0.08 (0.05) |
| Medial e' (m/s) | 0.07 (0.09) | 0.06 (0.02) |
| LAVI (mL/m<sup>2</sup>) | 42 (20) | 47 (21) |
| Peak TR Velocity (m/s) | 2.70 (0.38) | 2.85 (0.44) |
| PASP (mmHg) | 39 (12) | 42 (12) |
| *Quality of Life* |  |  |
| KCCQ (overall)* | 60 (19) | 54 (19) |
| KCCQ (clinical)* | 64 (17) | 55 (20) |
| MLHFQ<sup>†</sup> | 48 (23) | 44 (23) |
| NT-proBNP (pg/mL) | 678 (954) | 1001 (1128) |
| Hospitalization | 17 (19%) | 38 (17%) |
| Death | 1 (1.1%) | 3 (1.3%) |

Mean (SD); n (%)
Abbreviations: VO<sub>2</sub>, oxygen consumption; RER, respiratory exchange ratio; VE, minute ventilation; VCO<sub>2</sub>, carbon dioxide production; VE/VCO<sub>2</sub>, ventilatory efficiency; 6MWT, six-minute walk test; HR, heart rate; LAVI, left atrial volume index; TR, tricuspid regurgitation; PASP, pulmonary artery systolic pressure; KCCQ, Kansas City Cardiomyopathy Questionnaire; MLHFQ, Minnesota Living with Heart Failure Questionnaire; NT-proBNP, N-terminal fragment of the prohormone natriuretic peptide; mL, milliliter; kg, kilogram; min, minute; m, meter; s, second; mmHg, millimeter Mercury; pg, picogram
\*INDIE-HFpEF only
<sup>†</sup>RELAX only

### Echocardiographic Parameters

For echocardiographic parameters of diastolic function, HFpEF patients on beta blockers had a mean average E/e’ of 16 (SD 9), mean lateral e’ of 0.08 m/s (SD 0.05), and a mean medial e’ of 0.06 m/s (SD 0.02). Mean LAVI was 47 mL/m^2^ (SD 21), mean peak TR velocity was 2.85 m/s (SD 0.44), and mean PASP was 42 mmHg (SD 12). HFpEF patients not on beta blockers had a mean average E/e’ of 14 (SD 8), mean lateral e’ of 0.09 m/s (SD 0.03), and mean medial e’ of 0.07 m/s (SD 0.09). Mean LAVI was 42 mL/m^2^ (SD 20), mean peak TR velocity was 2.70 m/s (SD 0.38), and mean PASP was 39 mmHg (SD 12). In both groups, there was echocardiographic evidence of diastolic dysfunction as evidenced by the elevated average E/e’, left atrial enlargement, and elevated pulmonary artery systolic pressure.

### Quality of Life

Health-related quality of life was evaluated by baseline responses to KCCQ and MLHFQ surveys. Patients enrolled in RELAX completed MLHFQ, while patients enrolled in INDIE-HFpEF completed KCCQ (overall and clinical). Numerically, a KCCQ score of less than 50 suggests poor to fair health (with scores less than 25 representing poor health status), while MLHFQ scores above 45 represent poor perceived health status.^18,19^ HFpEF patients on beta blockers had a mean MLHFQ score of 44 (SD 23), while HFpEF patients not on beta blockers had a mean MLHFQ score of 48 (SD 23). HFpEF patients on beta blockers had a mean KCCQ overall score of 54 (SD 19), while HFpEF patients not on beta blockers had a mean KCCQ overall score of 60 (SD 19). HFpEF patients on beta blockers had a mean KCCQ clinical score of 55 (SD 20), while HFpEF patients not on beta blockers had a mean KCCQ clinical score of 64 (SD 17).HFpEF patients on beta blockers “normal” MLHFQ and KCCQ (overall and clinical) scores, while HFpEF patients not on beta blockers had perceived poor health status based upon MLHFQ scores despite “normal” KCCQ (overall and clinical) scores.

### Laboratory Assessment & Clinical Outcomes

At baseline, HFpEF patients on beta blockers had a mean NT-proBNP of 1001 pg/mL (SD 1128) compared to a mean NT-proBNP of 678 pg/mL (SD 954) in HFpEF patients not on beta blockers. At the conclusion of trial follow-up periods, 38 (17%) HFpEF patients on beta blockers had a HF hospitalization while 17 (19%) HFpEF patients not on beta blockers had a HF hospitalization. Three (1.3%) HFpEF patients on beta blockers died during the time of trial follow-up period, compared to 1 (1.1%) HFpEF patient not on beta blocker. In both groups, baseline NT-proBNP levels were elevated with modest incidence of HF hospitalization and/or death during trial follow-up.

### Regression Analysis

Results of multivariate linear regression are shown in Table 3. After controlling for potentially confounding variables (see methods), there were no significant differences in exercise parameters when comparing HFpEF patients on beta blockers to those not on beta blockers [peak VO2: estimate 0.028 (95% CI −0.635, 0.692); peak RER: estimate −0.012 (95% CI −0.037, 0.012); log-transformed VE/VCO2: estimate −0.014 (95% CI −0.086, 0.057); 6MWT: estimate 19.353 (95% CI −12.258, 50.965)], aside from peak HR (estimate −7.689 (95% CI −13.211, −2.167)). Similarly, there were no significant differences in echocardiographic parameters of diastolic function [log-transformed E/e’: estimate 0.092 (95% CI −0.026, 0.209); lateral e’: estimate −0.007 (95% CI −0.014, 0.000); medial e’: estimate −0.002 (95% CI −0.007, 0.003); LAVI: estimate 0.021 (95% CI −0.076, 0.119); peak TR velocity: estimate 0.093 (95% CI −0.039, 0.225); PASP: estimate 1.718 (95% CI −1.951, 5.387)].

**Table 3.** Multivariate linear regression to assess for differences in outcomes between heart failure with preserved ejection fraction patients taking beta blockers and those not taking beta blockers.

| Outcome | Estimate (95% Confidence Interval) | P Value |
| --- | --- | --- |
| *Exercise Parameters* |  |  |
| Peak VO<sub>2</sub> (mL/kg/min) | 0.028 (-0.635, 0.692) | 0.933 |
| Peak RER | -0.012 (-0.037, 0.012) | 0.315 |
| VE/VCO<sub>2</sub>, log-transformed* | -0.014 (-0.086, 0.057) | 0.687 |
| 6MWT (m)<sup>†</sup> | 19.353 (-12.258, 50.965) | 0.229 |
| Peak HR | -7.689 (-13.211, -2.167) | 0.007 |
| *Echocardiographic Parameters* |  |  |
| Average E/e', log-transformed | 0.092 (-0.026, 0.209) | 0.125 |
| Lateral e' (m/s) | -0.007 (-0.014, 0.000) | 0.055 |
| Medial e' (m/s) | -0.002 (-0.007, 0.003) | 0.468 |
| LAVI (mL/m<sup>2</sup>), log-transformed | 0.021 (-0.076, 0.119) | 0.665 |
| Peak TR Velocity (m/s) | 0.093 (-0.039, 0.225) | 0.165 |
| PASP (mmHg) | 1.718 (-1.951, 5.387) | 0.357 |
| *Symptom Assessments* |  |  |
| KCCQ (overall)* | -4.549 (-11.648, 2.550) | 0.206 |
| KCCQ (clinical)* | -6.542 (-13.654, 0.571) | 0.071 |
| MLHFQ<sup>†</sup> | -0.989 (-7.418, 5.440) | 0.762 |
| NT-proBNP (pg/mL), square-root | 1.948 (-1.087, 4.983) | 0.208 |

Abbreviations: VO<sub>2</sub>, oxygen consumption; RER, respiratory exchange ratio; VE, minute ventilation; VCO<sub>2</sub>, carbon dioxide production; VE/VCO<sub>2</sub>, ventilatory efficiency; 6MWT, six-minute walk test; HR, heart rate; LAVI, left atrial volume index; TR, tricuspid regurgitation; PASP, pulmonary artery systolic pressure; KCCQ, Kansas City Cardiomyopathy Questionnaire; MLHFQ, Minnesota Living with Heart Failure Questionnaire; NT-proBNP, N-terminal fragment of the prohormone natriuretic peptide; mL, milliliter; kg, kilogram; min, minute; m, meter; s, second; mmHg, millimeter Mercury; pg, picogram
\*INDIE-HFpEF only
<sup>†</sup>RELAX only

After multivariate linear regression, there were no significant differences in baseline quality of life survey scores when comparing HFpEF patients on beta blockers to those not on beta blockers [KCCQ (overall): estimate −4.549 (95% CI −11.648, 2.550); KCCQ (clinical): estimate - 6.542 (−13.654, 0.571); MLHFQ: estimate −0.989 (−7.418, 4.983)]. There was no significant difference in baseline NT-proBNP (square-root) after multivariate linear regression between HFpEF patients taking beta blockers compared to those not taking beta blockers [estimate 1.948 (95% CI −1.087, 4.983)]. Of note, clinical events (HF hospitalization and death) were deemed too infrequent to evaluate for statistically significant differences.

## DISCUSSION

In this secondary analysis of patient-level data from the INDIE-HFpEF and RELAX trials, there was no association with beta blocker use and worse exercise capacity, echocardiographic evidence of diastolic dysfunction, or quality of life in HFpEF patients. These results are somewhat contrary to our conventional understanding of the effects of beta blockers on exercise testing in HF patients, as prior publications have suggested using lower peak VO2 thresholds for risk stratification in patients tolerating beta blockers.^20^ Based upon mean values, both patient cohorts in this study had impaired baseline exercise capacity, with echocardiographic parameters and NT-proBNP levels suggestive of diastolic dysfunction. It is possible that a patient population with better baseline functional capacity may be more sensitive to the hemodynamic changes associated with beta blocker use, and that a healthier HFpEF phenotype may have demonstrated an association of beta blocker use with adverse exercise outcomes.

The baseline differences in medications between HFpEF patients taking beta blockers, compared to those who are not, is a point of interest. First, HFpEF patients on beta blockers were statistically more likely to also be taking a RAASi at baseline. This could be related to the observation that HFpEF patients on beta blockers were more likely to have hypertension, or a reflection of increased utilization of medications deemed beneficial for HF in this population. HFpEF patients on beta blockers were also more likely to be taking loop diuretics, compared to those not on beta blockers. This may also serve as an additional anti-hypertensive agent in this cohort, though mechanistically this may suggest that patients on beta blockers are more predisposed to increased filling pressures and volume overload. The association with increased utilization of these two medication classes in HFpEF patients taking beta blockers, compared to those who are not, has been observed in previous studies as well.^21^ HFpEF patients on beta blockers were also more likely to be on statins, despite no statistical difference in rate of prior MI between groups, though rates of coronary artery disease (without prior MI) was not assessed in this study.

HFpEF is a heterogenous disease process that encompasses multiple different phenotypes of patients with unique clinical features, comorbidities, and underlying pathophysiology.^22,23^ It is likely that HFpEF patients with different phenotypes, such as those with predominantly hypertensive heart disease, amyloidosis, hypertrophic cardiomyopathy, sarcoidosis, and beyond may respond differently to various interventions. It is possible that, in specific HFpEF patient cohorts, beta blockers have a substantial physiologic impact. Whether this impact results in meaningful changes in functional status likely depends on the HFpEF phenotype. Beyond altering physiologic or hemodynamic mechanisms, it remains unclear what clinical ramifications exist for beta blocker use in HFpEF patients, if any.

This rationale for differential treatment effect of beta blockers in HFpEF is supported by a recent post-hoc analysis of PRESERVE-HR, which demonstrated variations in response to beta blocker withdrawal according to LVEF and left ventricular size. Patients with lower baseline indexed LV end systolic volume had more robust increases in peak VO2, with trends for individuals with higher LVEF and smaller LV end diastolic volumes eliciting more benefit in terms of functional capacity with beta blocker withdrawal.^24^ This suggests that HFpEF patients with a limited stroke volume reserve may benefit more from an ability to augment HR in times of stress or exertion. Despite the lower resting and peak HR seen in the cohort of HFpEF patients taking beta blockers, indicative of some degree of therapeutic response to this medication class, exercise parameters were otherwise not significantly different between the two groups. This may be due to insufficient cohort size to detect mild differences in exercise parameters, or perhaps suggest that there are other mechanisms (independent of HR effects) by which beta blockers influence oxygen consumption.

Another plausible explanation for potential adverse effects of beta blockers in HFpEF patients, beyond impaired chronotropic response, is the negative inotropic effect of the medication class. In a randomized controlled trial comparing digoxin (which has neutral to mildly positive inotropic properties) to bisoprolol in older patients with permanent atrial fibrillation, there was no significant difference in the primary outcome of quality of life at 6 months. However, patients in the digoxin group appeared to have improved functional outcomes and reduced NT-proBNP levels, compared to increased NT-pro BNP levels in the bisoprolol group. Beyond this, there were significantly less treatment-related adverse effects in the digoxin group.^25^ This may be explained, at least in part, by the negative inotropic effect of beta blockers compared to the positive inotropic effect of digoxin.

Furthermore, there are a host of common comorbidities that exist in the HFpEF population that are classically managed with beta blockers, which include hypertension, atrial fibrillation, and coronary artery disease. However, the utility of beta blockers for these comorbidities, particularly in hypertension and chronic coronary artery disease, may not be as clinically important as previously believed. In a recently published HFpEF expert consensus decision pathway document, the authors recommend against the use of beta blockers for the management of hypertension due to reduced tolerability of beta blockers in HFpEF patients. Instead, loop diuretics, RAASi, and mineralocorticoid receptor antagonists are preferred.^26^ Notably, best practices for managing atrial fibrillation and coronary artery disease are currently being evaluated.

Regarding atrial fibrillation, beta blockers and non-dihydropyridine calcium channel blockers remain as mainstay therapies for patients deemed appropriate for a rate control strategy. However, most recent guidelines place a larger emphasis on a rhythm control strategy for HF patients, though these recommendations are largely supported by data obtained from studies of patients with HF with reduced ejection fraction. Despite the relative paucity of evidence, guidelines state that catheter ablation may improve quality of life and reduce symptoms in appropriate patients with HFpEF and atrial fibrillation.^27^ A review of patient outcomes from the Get with the Guidelines Heart Failure data registry demonstrated an association between rhythm control for atrial fibrillation with a lower risk of all-cause mortality in patients with HFPEF, compared to rate control.^28^

Several ongoing clinical trials are designed to better define the efficacy, safety, and quality of life associated with beta blocker use in patients with chronic coronary disease, including those with prior myocardial infarction without left ventricular systolic dysfunction.^29^ One recently published randomized clinical trial demonstrated no risk reduction for the composite primary endpoint of all-cause death or new myocardial infarction among patients with acute myocardial infarction who underwent early coronary angiography and had preserved LVEF with long-term beta blocker treatment.^30^ For patients with microvascular coronary dysfunction, which is common in HFpEF patients, current guidelines recommend beta blockers as first line therapy for angina.^31^ As most upcoming clinical trials will examine the effects of beta blockers in patients with coronary artery disease (including those with prior myocardial infarction) in patients with normal LVEF and no HF symptoms, some extrapolation will be required in translating these results to HFpEF patients.

This study has limitations that merit discussion. First, this was a retrospective analysis of prior randomized controlled trial data for clinical trials that were not designed to evaluate the effect of beta blockers. After creating two patient cohorts based upon beta blocker use, there were significant differences in baseline demographics. Patients enrolled in these trials were also not utilizing sodium-glucose cotransporter-2 inhibitors, which is now a guideline-recommended therapy for patients with HFpEF to decreased HF hospitalizations and cardiovascular mortality.^6^ Though attempts were made to control for confounding using multivariate linear regression, it is possible that other confounding factors were present and not accounted for in the statistical analysis. Though both studies were relatively small in terms of enrolled patients, the cohorts were combined in an attempt to maximize the potential to detect significant differences in outcomes. It is likely that an analysis of 320 patients was not adequate to identify subtle differences in outcomes between these populations, however, this study to assess beta blocker effects on exercise, diastolic function, and quality of life outcomes in HFpEF patients is currently the largest of its kind.

This study was also limited by a non-insignificant degree of missing data (missing outcomes data shown in Supplementary Table 2), particularly in terms of echocardiogram measurements. It is unclear why some patients did not have complete echocardiogram measurements, though it is possible that comprehensive evaluation of diastolic function was not able to be performed in patients in atrial fibrillation. Furthermore, some outcomes were assessed in only one of the two clinical trials. Similar to missing data, the smaller number of available patients for analysis of these outcomes diminishes the ability to detect mild differences. The outcomes for which data was available from only one trial (VE/VCO2, 6MWT, KCCQ, and MLHFQ) were determined to be important despite the relatively smaller population of patients evaluated, thus they were included in the final analysis. Finally, the clinical indication for beta blocker use, as well as dosing, in each patient included in this study is not known. However, it is unlikely that this population of patients were prescribed beta blockers solely for the management of HFpEF. Similarly, the length of time that each patient was taking beta blockers prior to trial enrollment was also unable to be determined based upon available data.

This study adds further insight into our current understanding of the effects, or lack thereof, of beta blockers in patients with HFpEF. Based on the results of this study, beta blockers do not appear to influence exercise capacity, diastolic function, or quality of life in HFpEF patients. In the contemporary management of patients with heart failure with preserved ejection fraction, there likely is a cohort of patients that may derive benefit from beta blocker administration. However, implementation of beta blockers in HFpEF patients requires nuance and thoughtful consideration for the underlying pathophysiology and clinical presentation. With alternative medication classes available for the management of atrial fibrillation, angina secondary to coronary artery disease, and hypertension, it may be reasonable to trial discontinuation of beta blockers in patients who appear to have adverse effects from this medication class. Recruiting and examining patients with distinct HFpEF phenotypes will be paramount in effectively evaluating the impact of different interventions, including the use of beta blockers, and improving our understanding of the pathophysiology of HFpEF.

This study demonstrated no significant association between beta blocker use and worsened exercise capacity, echocardiographic evidence of diastolic dysfunction, or quality of life in HFpEF patients. Given the lack of proven clinical or symptomatic benefit of beta blocker use in HFpEF patients, along with improvements in exercise capacity after beta blocker withdrawal in select HFpEF patients, further evaluation is warranted with randomized controlled trials. It is necessary to delineate HFpEF phenotypes to maximize the utility of future clinical trials, and to correctly apply trial results to appropriate populations of patients.

## Data Availability

Deidentified patient-level data for this study was obtained via request from the NIH National Heart, Lung, and Blood Institute Biologic Specimen and Data Repository Information Coordinating Center.

https://biolincc.nhlbi.nih.gov/studies/hfn_relax/

https://biolincc.nhlbi.nih.gov/studies/hfn_indie/

## NONSTANDARD ABBREVIATIONS

INDIE-HFpEF: Effect of Inorganic Nitrite vs Placebo on Exercise Capacity Among Patients with Heart Failure with Preserved Ejection Fraction;
PRESERVE-HR: Effect of β-Blocker Withdrawal on Functional Capacity in Heart Failure and Preserved Ejection Fraction;
RELAX: Effect of phosphodiesterase-5 inhibition on exercise capacity and clinical status in heart failure with preserved ejection fraction: a randomized clinical trial;
TOPCAT: Spironolactone for heart failure with preserved ejection fraction

## ACKNOWLEDGEMENTS

The authors gratefully acknowledge the contributions of the Ohio State University Center for Biostatistics, with special thanks to Jianing Ma, MS and Jing Peng, PhD.

## SUPPLEMENTAL MATERIAL

Tables S1-S2

## REFERENCES

1. Bozkurt B, Ahmad T, Alexander KM, et al. Heart Failure Epidemiology and Outcomes Statistics: A Report of the Heart Failure Society of America. J Card Fail. 2023;29:1412– 1451.

2. Pitt B, Pfeffer MA, Assmann SF, et al. Spironolactone for heart failure with preserved ejection fraction. N Engl J Med. 2014;370:1383–92.

3. Solomon SD, McMurray JJ V, Anand IS, et al. Angiotensin-Neprilysin Inhibition in Heart Failure with Preserved Ejection Fraction. N Engl J Med. 2019;381:1609–1620.

4. Anker SD, Butler J, Filippatos G, et al. Empagliflozin in Heart Failure with a Preserved Ejection Fraction. N Engl J Med. 2021;385:1451–1461.

5. Solomon SD, McMurray JJ V, Claggett B, et al. Dapagliflozin in Heart Failure with Mildly Reduced or Preserved Ejection Fraction. N Engl J Med. 2022;387:1089–1098.

6. Heidenreich PA, Bozkurt B, Aguilar D, et al. 2022 AHA/ACC/HFSA Guideline for the Management of Heart Failure: Executive Summary: A Report of the American College of Cardiology/American Heart Association Joint Committee on Clinical Practice Guidelines. J Am Coll Cardiol. 2022;79:1757–1780.

7. Xu X, Wang DW. The progress and controversial of the use of beta blockers in patients with heart failure with a preserved ejection fraction. IJC Hear Vasc. 2020;26:100451.

8. Wernhart S, Papathanasiou M, Rassaf T, Luedike P. The controversial role of beta-blockers in heart failure with preserved ejection fraction. Pharmacol Ther. 2023;243:108356.

9. Fudim M, Kaye DM, Borlaug BA, et al. Venous Tone and Stressed Blood Volume in Heart Failure: JACC Review Topic of the Week. J Am Coll Cardiol. 2022;79:1858–1869.

10. Yamamoto K, Origasa H, Hori M, J-DHF Investigators. Effects of carvedilol on heart failure with preserved ejection fraction: the Japanese Diastolic Heart Failure Study (J-DHF). Eur J Heart Fail. 2013;15:110–8.

11. Zafeiropoulos S, Farmakis IT, Milioglou I, et al. Pharmacological Treatments in Heart Failure with Mildly Reduced and Preserved Ejection Fraction: Systematic Review and Network Meta-Analysis. JACC Heart Fail. 2023;S2213-1779(23)00410–9.

12. Arnold S V., Silverman DN, Gosch K, et al. Beta-Blocker Use and Heart Failure Outcomes in Mildly Reduced and Preserved Ejection Fraction. JACC Heart Fail. 2023;11:893–900.

13. Silverman DN, Plante TB, Infeld M, et al. Association of β-Blocker Use with Heart Failure Hospitalizations and Cardiovascular Disease Mortality Among Patients with Heart Failure With a Preserved Ejection Fraction: A Secondary Analysis of the TOPCAT Trial. JAMA Netw Open. 2019;2:e1916598.

14. Palau P, Seller J, Domínguez E, et al. Effect of β-Blocker Withdrawal on Functional Capacity in Heart Failure and Preserved Ejection Fraction. J Am Coll Cardiol. 2021;78:2042–2056.

15. Ross JS, Ritchie JD, Finn E, Desai NR, Lehman RL, Krumholz HM, Gross CP. Data sharing through an NIH central database repository: a cross-sectional survey of BioLINCC users. BMJ Open. 2016;6:e012769.

16. Borlaug BA, Anstrom KJ, Lewis GD, et al. Effect of Inorganic Nitrite vs Placebo on Exercise Capacity Among Patients with Heart Failure With Preserved Ejection Fraction: The INDIE-HFpEF Randomized Clinical Trial. JAMA. 2018;320:1764–1773.

17. Redfield MM, Chen HH, Borlaug BA, et al. Effect of phosphodiesterase-5 inhibition on exercise capacity and clinical status in heart failure with preserved ejection fraction: a randomized clinical trial. JAMA. 2013;309:1268–77.

18. Hsu T-W, Chang H-C, Huang C-H, Chou M-C, Yu Y-T, Lin L-Y. Identifying cut-off scores for interpretation of the Heart Failure Impact Questionnaire. Nurs Open. 2018;5:575–582.

19. Spertus JA, Jones PG, Sandhu AT, Arnold S V. Interpreting the Kansas City Cardiomyopathy Questionnaire in Clinical Trials and Clinical Care: JACC State-of-the-Art Review. J Am Coll Cardiol. 2020;76:2379–2390.

20. Malhotra R, Bakken K, D’Elia E, Lewis GD. Cardiopulmonary Exercise Testing in Heart Failure. JACC Heart Fail. 2016;4:607–16.

21. Formiga F, Chivite D, Nuñez J, et al. Beta-blocker use in patients with heart failure with preserved ejection fraction and sinus rhythm. Rev Port Cardiol. 2022;41:853–861.

22. Redfield MM, Borlaug BA. Heart Failure With Preserved Ejection Fraction: A Review. JAMA. 2023;329:827–838.

23. Shah SJ, Kitzman DW, Borlaug BA, et al. Phenotype-Specific Treatment of Heart Failure with Preserved Ejection Fraction: A Multiorgan Roadmap. Circulation. 2016;134:73–90.

24. Palau P, de la Espriella R, Seller J, et al. β-Blocker Withdrawal and Functional Capacity Improvement in Patients with Heart Failure With Preserved Ejection Fraction. JAMA Cardiol. 2024:1–5.

25. Kotecha D, Bunting KV, Gill SK, et al. Effect of Digoxin vs Bisoprolol for Heart Rate Control in Atrial Fibrillation on Patient-Reported Quality of Life: The RATE-AF Randomized Clinical Trial. JAMA. 2020;324(24):2497–2508.

26. Kittleson MM, Panjrath GS, Amancherla K, et al. 2023 ACC Expert Consensus Decision Pathway on Management of Heart Failure with Preserved Ejection Fraction: A Report of the American College of Cardiology Solution Set Oversight Committee. J Am Coll Cardiol. 2023;81:1835–1878.

27. Joglar JA, Chung MK, Armbruster AL, et al. 2023 ACC/AHA/ACCP/HRS Guideline for the Diagnosis and Management of Atrial Fibrillation: A Report of the American College of Cardiology/American Heart Association Joint Committee on Clinical Practice Guidelines. Circulation. 2024;149:e1–e156.

28. Kelly JP, DeVore AD, Wu J, et al. Rhythm Control Versus Rate Control in Patients With Atrial Fibrillation and Heart Failure With Preserved Ejection Fraction: Insights From Get With The Guidelines-Heart Failure. J Am Heart Assoc. 2019;8:e011560.

29. Steg PG. Routine Beta-Blockers in Secondary Prevention - On Injured Reserve. N Engl J Med. 2024;390:1434–1436.

30. Yndigegn T, Lindahl B, Mars K, et al. Beta-Blockers after Myocardial Infarction and Preserved Ejection Fraction. N Engl J Med. 2024:1–10.

31. Virani SS, Newby LK, Arnold S V., et al. 2023 AHA/ACC/ACCP/ASPC/NLA/PCNA Guideline for the Management of Patients with Chronic Coronary Disease: A Report of the American Heart Association/American College of Cardiology Joint Committee on Clinical Practice Guidelines. Circulation. 2023;148:e9–e119.

